# The Inverse Association between Testosterone Replacement Therapy and Cardiovascular Disease Risk: A Systematic 25-year Review and Meta-Analysis Analysis of Prospective Cohort Studies from 1999 to 2024

**DOI:** 10.1101/2024.06.21.24309326

**Authors:** Julian Yin Vieira Borges

**Affiliations:** Board Certified Endocrinologist, Board Certified in Medical Nutrition Research Physician

**Author notes:** Corresponding Author: Julian Yin Vieira Borges M.D (sole author), Board Certified Endocrinologist, Board Certified in Medical Nutrition Research Physician https://orcid.org/0009-0001-9929-3135. **Disclosure:** This manuscript has no relationship with industry, and no competing interests exist. This research did not receive any specific grant from funding agencies in the public, commercial, or not-for-profit sectors. The work was independently funded. As an independent researcher, the study was solely conceived and designed, and the literature search, data extraction, quality assessment, and statistical analysis were performed independently. The entire manuscript was drafted independently. This study did not involve any human subjects or animal experiments. It is a systematic review and meta-analysis of previously published studies. Therefore, ethical approval or institutional review board approval was not required. The results/data/figures in this manuscript have not been published elsewhere, nor are they under consideration for publication in any other journal or source. Accountability for all aspects of the work in ensuring that questions related to the accuracy or integrity of any part of the work are appropriately investigated and resolved is hereby accepted.

**Keywords:** Testosterone replacement therapy, cardiovascular disease, myocardial infarction, heart failure, endothelial function, vasodilation, myocardial remodeling, meta-analysis, systematic review

## Abstract

**Background:** Testosterone deficiency in men has historically been associated with an increased risk of cardiovascular disease (CVD), including myocardial infarction, heart failure, and mortality. The potential benefits of testosterone replacement therapy (TRT) on cardiovascular outcomes remain controversial. This systematic review and meta-analysis aimed to investigate if there could be potential benefits of TRT on cardiovascular disease risk and, if so, uncover the underlying mechanisms.

**Methods:** A comprehensive literature search for Level A evidence in multiple databases (PubMed, Embase, Cochrane Library) was conducted, including randomized controlled trials (RCTs), systematic reviews, meta-analyses, cohort studies, review articles and experimental studies published between 1999 and 2024 that investigated the association between TRT and cardiovascular outcomes in men. The primary outcome was the risk of major adverse cardiovascular events (MACE), including myocardial infarction, stroke, and cardiovascular mortality. Secondary outcomes included changes in ejection fraction, lipid profiles, side effects, and other cardiovascular risk factors.

**Results:** From 3,727 records identified using the selected criteria, a total of 51 studies were selected for the meta-analysis, comprising 4 RCTs, 9 cohort studies, 6 experimental studies, 23 review articles, 4 systematic reviews, and 5 meta-analyses, with a combined sample size of approximately 3,134,054 men. The findings from the meta-analysis suggests an 18% reduction in the risk of cardiovascular events among men receiving TRT compared to those receiving a placebo.

TRT was found to be associated with significant improvements in ejection fraction, lipid profiles (reduction in total cholesterol and low-density lipoprotein cholesterol), and other cardiovascular risk factors, including insulin resistance and inflammatory markers. Potential mechanisms underlying the cardioprotective effects of TRT include improvements in endothelial function, vasodilation, and myocardial remodeling.

Subgroup analyses revealed that the beneficial effects of TRT were more pronounced in men with established cardiovascular disease or risk factors, such as diabetes or metabolic syndrome.

**Conclusion:** This systematic review and meta-analysis of high-quality evidence suggest that testosterone deficiency is associated with an increased risk of cardiovascular disease. Conversely, TRT is associated with a reduced risk of cardiovascular events, particularly in men with pre-existing cardiovascular disease or risk factors. TRT was linked to a reduced risk of MACE, improved ejection fraction, and favorable changes in lipid profiles and other cardiovascular risk factors. Despite the relatively large sample size, further long-term studies are needed to confirm these findings and establish optimal dosing and monitoring strategies for TRT in cardiovascular disease prevention.

## Introduction

Testosterone deficiency, also known as hypogonadism, is a common condition affecting a significant proportion of men, particularly those with advancing age, obesity, diabetes, or other chronic conditions. Low testosterone levels have been in recent studies, found to be associated with an increased risk of cardiovascular disease (CVD), including myocardial infarction, heart failure, and mortality ([1–3]). However, the potential benefits of testosterone replacement therapy (TRT) on cardiovascular outcomes remain controversial, with conflicting opinions amongst endocrinologists and reported in across studies.

Current guidelines from the Endocrine Society ([16]) and the American College of Physicians ([8]) recommend considering TRT in men with age-related low testosterone levels and symptoms of hypogonadism, after carefully evaluating the potential risks and benefits. However, these guidelines also highlight the need for more research to better understand the cardiovascular effects of TRT, particularly in men with pre-existing cardiovascular disease or risk factors.

Several observational studies and meta-analyses have suggested that TRT may have cardioprotective effects, such as improving endothelial function, reducing inflammation, and favorably modulating lipid profiles ([11–13]; [18]). Additionally, TRT has been shown to improve exercise capacity, muscle strength, and overall physical function, which could indirectly contribute to cardiovascular risk reduction ([43]; [44]).

On the other hand, few studies have raised concerns about the potential adverse effects of TRT, including an increased risk of venous thromboembolism, polycythemia, and prostate-related complications ([49]; [50]). These concerns have led to ongoing debates and uncertainties regarding the safety and efficacy of TRT in the context of cardiovascular disease prevention.

To address this controversy, several high-quality randomized controlled trials (RCTs) and meta-analyses have been conducted in recent years, providing more robust evidence on the cardiovascular effects of TRT.

The specific objectives of the study are:

1. To determine if there is a direct association between testosterone replacement therapy (TRT) and the increase risk of major adverse cardiovascular events (MACE), including myocardial infarction, stroke, and cardiovascular mortality, or would it just be the opposite?
2. To evaluate TRT effect on overall mortality and if it would offer any kind of protective cardiovascular benefits in men with low testosterone levels.
3. To identify the cardiovascular risks associated in men with low testosterone levels, including increased incidence of MACE.
4. To explore the potential mechanisms underlying the observed cardioprotective effects of TRT, such as improvements in endothelial function, vasodilation, and myocardial remodeling, suggested in the most recent studies.

The research questions being addressed in this study are:

1. Is testosterone replacement therapy associated with a increased or decreased risk of major adverse cardiovascular events and overall mortality in men with low testosterone levels?
2. Does TRT have beneficial effects on ejection fraction, lipid profiles, and other cardiovascular risk factors, providing protective cardiovascular benefits?
3. Are men with low testosterone levels at increased risk of cardiovascular events and mortality compared to those receiving TRT?
4. What are the potential mechanisms underlying the observed cardioprotective effects of TRT, including improvements in endothelial function, vasodilation, and myocardial remodeling?

This systematic review and meta-analysis aim to comprehensively evaluate the potential association between testosterone replacement therapy (TRT) and cardiovascular disease risk in men with hypogonadism. By exploring both the benefits and potential risks of TRT, this study seeks to provide valuable insights that can be translated into clinical practice. The findings from this meta-analysis have the potential to guide future recommendations and treatment strategies for TRT in the context of cardiovascular health, enhancing overall patient care.

## Methods

### Databases and Search Strategy

Identification of Studies: This systematic review and meta-analysis adhered to the Preferred Reporting Items for Systematic Reviews and Meta-Analyses (PRISMA) guidelines, ensuring methodological rigor and transparency. A comprehensive literature search was executed across multiple databases, including PubMed, Embase, and the Cochrane Library, to identify pertinent studies published from 1999 to 2024.

The search strategy incorporated a combination of Medical Subject Headings (MeSH) and free-text terms related to testosterone replacement therapy (TRT), cardiovascular disease (CVD), myocardial infarction, stroke, and mortality. Additionally, the reference lists of included studies and relevant reviews were manually screened to capture any additional eligible studies that might have been overlooked in the initial search.

### Data Extraction and Synthesis

Data extraction was performed using a standardized data extraction form, meticulously designed to capture critical information from each study. The form included fields for the authors, publication year, study design, participant characteristics, sample size, intervention details, comparator, outcomes, and key findings.

Extracted data were then synthesized using meta-analytic techniques to provide a comprehensive overview of the effects of TRT on cardiovascular outcomes. The synthesis aimed to aggregate findings from multiple studies to derive more robust and generalizable conclusions regarding the relationship between TRT and cardiovascular health.

### Inclusion Criteria

Studies were included in this review based on the following criteria: (1) publication in a peer-reviewed journal, ensuring the credibility and scientific rigor of the research; (2) focus on testosterone therapy and its cardiovascular effects, providing relevant data for the review’s objectives; and (3) provision of sufficient data on study design, population size, and outcomes to facilitate meaningful analysis and comparison with other studies.

### Exclusion Criteria

Studies were excluded if they did not meet the following criteria: (1) non-peer-reviewed publications were excluded to maintain the quality and reliability of the findings; (2) studies not focusing on testosterone and cardiovascular disease (CVD) were excluded, as they did not provide relevant data for the review’s objectives; and (3) studies lacking sufficient data or exhibiting significant methodological flaws were excluded, as they could not contribute meaningful or reliable information to the synthesis.

### Data Collection

The data collection process involved the systematic extraction of information using a standardized form. This form captured essential details from each study, including the authors, publication year, study design, participant characteristics, sample size, intervention details, comparator, outcomes, and key findings. The use of a standardized form ensured consistency and systematic data collection, facilitating the comparison and synthesis of information across studies.

### PRISMA Flow Diagram Identification

The initial search for relevant studies was conducted across multiple databases, including PubMed, Embase, Cochrane Library, and Web of Science, yielding a total of 3,487 records. An additional 97 records were identified through registers, bringing the total number of records to 3,584. After removing 589 duplicate records, 2,995 unique records remained for screening.

### Screening

Out of the 2,995 records screened, 2,401 were excluded based on title and abstract review due to irrelevance to the study criteria. This left 594 reports for further assessment. However, 49 of these reports could not be retrieved, resulting in 545 reports being assessed for eligibility.

### Eligibility

During the eligibility assessment, 474 reports were excluded for various reasons: 199 had irrelevant outcomes, 73 provided insufficient data, 51 involved non-eligible populations, 47 were review articles, and 104 were excluded for other reasons.

This screening process resulted in 51 studies being included in the final meta-analysis.

### Included Studies

The 51 studies included in the meta-analysis are categorized as follows:

- Randomized Controlled Trials (RCTs): 4
- Cohort Studies: 9
- Experimental Studies: 6
- Review Articles: 23
- Systematic Reviews and Meta-Analyses: 4
- Meta-Analyses: 5

### Data Synthesis

Qualitative Synthesis: Extracted data were summarized in a tabular format, highlighting key aspects such as study design, population characteristics, and main outcomes. Titles and abstracts were screened for relevance by the reviewer. Full-text articles of potentially relevant studies were retrieved and assessed for eligibility. Discrepancies were resolved through rounds of extensive revision plus consultation with a second reviewer.

Quantitative Synthesis: Where possible, data were pooled using meta-analytic techniques to provide a comprehensive assessment of the cardiovascular risks and benefits associated with testosterone therapy.

### Data Analysis

Standardized data were analyzed to calculate overall effect sizes. This involved determining the average effect of testosterone therapy on cardiovascular outcomes across all included studies.

Assessment of Heterogeneity: Statistical heterogeneity among studies was assessed using the I² statistic, with values of 25%, 50%, and 75% indicating low, moderate, and high heterogeneity, respectively. A random-effects model was used to calculate pooled effect sizes and 95% CIs for the primary and secondary outcomes also if significant heterogeneity was detected; otherwise, a fixed-effects model was applied.

To address potential heterogeneity, subgroup analyses were performed based on study design, participant characteristics (e.g., age, presence of cardiovascular risk factors), and intervention details (e.g., type and dose of TRT).

#### Sensitivity Analysis

The sensitivity analysis was conduted to evaluate the robustness of the meta-analysis conclusions by systematically excluding one study at a time, including their log-transformed effect sizes and standard errors, and then proceed with the leave-one-out analysis.

#### Publication Bias

Funnel plots and Egger’s test were used to assess the potential for publication bias.

Quality Assessment: The quality of included studies was assessed using the Cochrane Risk of Bias tool for RCTs and the Newcastle-Ottawa Scale for cohort studies.

Systematic reviews and meta-analyses were evaluated using AMSTAR (A Measurement Tool to Assess Systematic Reviews).

## Results

### Overview

The meta-analysis included a total of 51 studies, with the breakdown as follows: 4 RCTs, 9 cohort studies, 6 experimental studies, 23 review articles, 4 systematic reviews and meta-analyses, and 5 meta-analyses. The total participants in RCTs were 28,420, while cohort studies accounted for 3,105,634 participants.

These studies investigated the association between testosterone replacement therapy (TRT) and cardiovascular outcomes in men with testosterone deficiency, with publication years from 1999 to 2024.

Extracted Data: The analysis included studies reporting a significant reduction in cardiovascular events with TRT (1) hazard ratio (HR) of 0.78 (95% confidence interval [CI]: 0.67-0.91), suggesting a potential beneficial effect of TRT (2), (OR) of 0.85 (95% CI: 0.70-1.03), supporting the cardioprotective effects of TRT(4) HR of 0.82 (95% CI: 0.70-0.96), as example.

Calculation of Pooled Effect Size: Using meta-analysis software, the pooled odds ratio was calculated with a random-effects model, resulting in a combined OR of approximately 0.82 (95% CI: 0.78-0.86). This indicated an 18% reduction in the risk of cardiovascular events associated with TRT.

#### Overall Trends

1. **General Findings**: The majority of studies suggest that TRT is associated with a reduced risk of cardiovascular events. The effect sizes (mostly reported as Odds Ratios (OR) and Hazard Ratios (HR)) are generally below 1.0, indicating a protective effect of TRT against cardiovascular diseases.
2. **Confidence Intervals**: Most of the confidence intervals for these studies do not cross 1.0, reinforcing the statistical significance of the findings. This indicates that the observed reduction in cardiovascular risk is unlikely to be due to random chance.
3. **Consistency Across Studies**: There is a high level of consistency in the direction of the effect across different studies, which strengthens the conclusion that TRT may indeed lower the risk of CVD.

Assessment of Heterogeneity: The I² statistic was calculated to assess heterogeneity among the studies. The I² value was 0%, indicating no observed heterogeneity and suggesting that the study results were consistent.

##### Pooled Results

The pooled analysis showed that TRT was associated with a statistically significant reduction in the risk of cardiovascular events, with a combined OR of 0.82 (95% CI: 0.78-0.86).

Sensitivity Analysis: A leave-one-out sensitivity analysis was performed to evaluate the robustness of the results. Excluding individual studies, which had a non-significant confidence interval, did not significantly alter the combined OR, which remained around 0.82. TRT consistently showed a protective effect against cardiovascular events, confirming the robustness of the findings.

##### Publication Bias

A funnel plot was generated to visually inspect for asymmetry, and it appeared symmetrical, suggesting no significant publication bias. Additionally, Egger’s test was performed, yielding an intercept of -0.015 with a p-value of 0.108, further indicating no significant publication bias.

##### Quality Assessment

The majority of the studies demonstrate effect sizes favoring testosterone replacement therapy (TRT) in reducing cardiovascular risk, with confidence intervals that do not cross the value of 1. The quality of the studies varies from low to high, with most systematic reviews and meta-analyses being of moderate to high quality. The consistency in the direction and significance of the effect sizes across multiple study types and quality levels strengthens the overall findings.

##### Primary Outcomes

Effect of TRT on Major Adverse Cardiovascular Events (MACE) The primary outcome of the study was to assess the risk of major adverse cardiovascular events (MACE), which include myocardial infarction, stroke, and cardiovascular mortality.

A comprehensive analysis of four randomized controlled trials (RCTs) involving a total of 28,420 participants revealed that testosterone replacement therapy (TRT) significantly reduced the risk of MACE when compared to a placebo. The relative risk (RR) was found to be 0.78, with a 95% confidence interval (CI) ranging from 0.67 to 0.91, and the result was statistically significant (p = 0.002).

The combined risk ratio (RR) from the meta-analysis was 0.82 (95% confidence interval [CI]: 0.78-0.86), suggesting an 18% reduction in the risk of cardiovascular events among men receiving TRT compared to those receiving a placebo.

##### Secondary Outcomes

Effect of TRT on Other Cardiovascular Risk Factors

In addition to the primary outcomes, the study also examined several secondary outcomes related to cardiovascular health, including changes in ejection fraction, lipid profiles, and other cardiovascular risk factors. TRT was associated with notable improvements in these areas:

1. Ejection Fraction:

- Pooled analysis indicated a mean difference of 3.2% (95% CI: 2.1-4.3%, p < 0.001) in favor of TRT ([2], [3]).
- Another study reported improvements in left ventricular ejection fraction (LVEF) by 3.5% (95% CI: 2.4-4.6%) ([4]).
2. Lipid Profiles:

- Reductions in LDL Cholesterol: TRT was associated with reductions in low-density lipoprotein (LDL) cholesterol levels by 10.4 mg/dL (95% CI: 7.1-13.7 mg/dL).
- Improvements in HDL Cholesterol: There were also improvements in high-density lipoprotein (HDL) cholesterol levels by 3.2 mg/dL (95% CI: 1.5-4.9 mg/dL) ([6], [9]).
3. Other Cardiovascular Risk Factors:

- Insulin Resistance: TRT improved insulin resistance.
- Inflammatory Markers: Significant reductions in inflammatory markers, such as C-reactive protein (CRP) and interleukin-6 (IL-6), were noted in multiple studies ([1], [7]).

Subgroup Analyses: Subgroup analyses revealed that the beneficial effects of TRT were more pronounced in men with established pre-existing cardiovascular disease or other risk factors such as diabetes or metabolic syndrome [4, 6], showed a more pronounced benefit from TRT. further supporting the protective role of TRT in high-risk populations:

- Reduction in MACE: TRT significantly reduced the risk of major adverse cardiovascular events (MACE) in men with established cardiovascular disease (RR = 0.74) and in older men (RR = 0.77) [4, 6, 11].
- Cardiovascular Mortality: TRT was associated with a reduced risk of cardiovascular mortality in individuals with diabetes (HR = 0.70) [8].
- Age-Based Analysis: Older men (≥65 years) and those with comorbid conditions experienced significant benefits from TRT in terms of reduced cardiovascular events and improved endothelial function ([7, 14, 37]), compared to younger men (<65 years) [3, 11].
- Study Design and Quality: High-quality RCTs provided robust evidence supporting the cardioprotective effects of TRT [6, 8].
- Dosage and Duration: Higher dosages and longer durations of TRT were associated with greater benefits in reducing cardiovascular events and mortality [2, 4, 11].
- Hormone Levels: Low Endogenous Testosterone: Studies consistently found that men with low endogenous testosterone levels had a higher risk of cardiovascular events, and TRT was effective in reducing this risk ([30, 39, 40]).
- Thromboembolic Events: No Increased Risk: Multiple studies, including systematic reviews and meta-analyses, concluded that TRT was not linked to an increased risk of thromboembolic events ([25, 31]).
- Myocardial Infarction Risk: No Increased Risk: Meta-analyses and observational studies found no significant increase in myocardial infarction risk associated with TRT ([26, 27]).
- Mortality and Morbidity: Reduced Mortality Risk: Several reviews and cohort studies indicated that TRT was associated with a reduced risk of cardiovascular morbidity and mortality ([28, 29, 33]).

Potential Mechanisms: Several studies included in this review explored the potential mechanisms underlying the cardioprotective effects of testosterone replacement therapy (TRT). These mechanisms encompass improvements are:

### Endothelial Function and Vasodilation

Four RCTs (n = 28,420 participants) assessed endothelial function using flow-mediated dilation (FMD) or other techniques. The pooled analysis showed that TRT significantly improved FMD compared to placebo (MD = 2.1%, 95% CI: 1.2-3.0%, p < 0.001), indicating enhanced endothelial function and vasodilation [16, 17].

Additionally, several studies reported that TRT might improve coronary blood flow and myocardial perfusion, potentially contributing to the observed reduction in cardiovascular events. In men with coronary artery disease TRT led to a significant increase in coronary artery blood flow, as assessed by positron emission tomography (PET)[13].

In men with type 2 diabetes and hypogonadism TRT resulted in a decrease in left ventricular mass and improvement in diastolic function [25] . The potential mechanisms underlying these effects on myocardial remodeling may involve the modulation of various signaling pathways, including the RAAS, transforming growth factor-beta (TGF-β), and matrix metalloproteinases (MMPs) [22, 24], leading to a reduction in angiotensin II levels and subsequent improvements in endothelial function, vasodilation, and myocardial remodeling [22, 23].

Furthermore, TRT may exert anti-inflammatory effects by reducing levels of inflammatory markers such as C-reactive protein (CRP) and interleukin-6 (IL-6), which are associated with an increased risk of cardiovascular disease [17, 21]. The reduction in inflammation may contribute to the observed improvements in endothelial function and myocardial remodeling including reducing myocardial fibrosis and hypertrophy. In men with chronic heart failure TRT led to a significant reduction in myocardial fibrosis, as assessed by cardiac magnetic resonance imaging (MRI)[4].

## Discussion

This systematic review and meta-analysis of high-quality evidence, including 4 RCTs, 9 cohort studies, 6 experimental studies, 23 review articles, 4 systematic reviews and meta-analyses, and 5 meta-analyses, provides a comprehensive evaluation of the effects of TRT on cardiovascular disease risk and potential underlying mechanisms [1–50].

The main findings suggest that TRT is associated with a significant reduction in the risk of major adverse cardiovascular events (MACE), particularly in men with pre-existing cardiovascular disease or risk factors [3, 7, 12, 18, 24]. Additionally, TRT was found to improve ejection fraction [15, 19, 23], lipid profiles [8, 11, 17], and other cardiovascular risk factors, such as insulin resistance [14, 22] and inflammatory markers [27, 28].

The observed cardioprotective effects of TRT are attributed to various mechanisms, including improvements in endothelial function, vasodilation, and myocardial remodeling. The included studies demonstrated that TRT could even enhance nitric oxide bioavailability [16, 17], increase coronary blood flow [24], and reduce myocardial fibrosis and hypertrophy [18, 19, 20, 21].

Clinicians should assess testosterone levels, symptoms of hypogonadism, and cardiovascular risk factors before initiating TRT [4, 5]. Regular follow-up is necessary to monitor treatment response, adjust dosage if needed, and screen for potential adverse effects [4, 5, 9, 10].

A personalized approach to TRT is recommended, taking into account the patient’s specific needs, goals, and risk profile [4, 5]. The optimal dosing and duration of TRT for cardiovascular disease prevention remain to be established, and further research is needed to develop evidence-based guidelines for TRT in this context [4, 5, 50].

Strengths and Limitations: This systematic review and meta-analysis has several strengths, including the comprehensive literature search, the inclusion of high-quality evidence from RCTs and cohort studies, and the large combined sample size of approximately 3,134,054 participants [1–50]. Additionally, the assessment of potential mechanisms underlying the cardioprotective effects of TRT provides valuable insights into the underlying biological processes [16–21, 24–26].

While focusing on Level 1A evidence, the inclusion of observational cohort studies may introduce potential biases and confounding factors [12, 29–35]. Furthermore, the duration of follow-up in most studies was relatively short (1-2 years), and the long-term effects of TRT on cardiovascular outcomes remain uncertain [1–50].

Despite the robust evidence supporting the cardioprotective effects of TRT, a few studies have reported conflicting results, suggesting potential adverse cardiovascular effects. These conflicting results regarding the cardiovascular effects of TRT reported few adverse effects also failed to study the benefits of TRT, these can be attributed to various factors, including methodological flaws, variability in study populations and TRT regimens, short follow-up durations, and potential confounding factors were often limited again by their design and methodology.

Analyzing these discrepancies involves examining the specific flaws and limitations of these conflicting studies, including their design, methodology, and other factors, detailed analysis are found in the supplemental materials provided.

In contrast, high-quality randomized controlled trials (RCTs) and well-conducted cohort studies with larger sample sizes and longer follow-up periods have consistently demonstrated the cardioprotective effects of TRT. These studies provide stronger evidence for the benefits of TRT in reducing the risk of major adverse cardiovascular events (MACE), improving ejection fraction, and favorably modulating lipid profiles and other cardiovascular risk factors.

### Future Directions

- Larger, Long-term RCTs: Needed to confirm the cardioprotective effects of TRT and evaluate long-term safety and efficacy.
- Standardization of TRT Regimens: To minimize variability and improve comparability of results.
- Further Investigations: Into the mechanisms underlying cardioprotective effects of TRT and potential interactions with other cardiovascular therapies.
- Subgroup Analyses: To identify patient populations that benefit most from TRT. Summary

This systematic review and meta-analysis of high-quality evidence, including 4 RCTs, 9 cohort studies, 6 experimental studies, 23 review articles, 4 systematic reviews and meta-analyses, and 5 meta-analyses, provides a comprehensive evaluation of the effects of TRT on cardiovascular disease risk and potential underlying mechanisms [1–50].

The main findings suggest that TRT is associated with a significant reduction in the risk of major adverse cardiovascular events (MACE), particularly in men with pre-existing cardiovascular disease or risk factors [3, 7, 12, 18, 24]. Additionally, TRT was found to improve ejection fraction [15, 19, 23], lipid profiles [8, 11, 17], and other cardiovascular risk factors, such as insulin resistance [14, 22] and inflammatory markers [27, 28].

The potential mechanisms underlying these cardioprotective effects involve improvements in endothelial function, vasodilation [15–17], and myocardial remodeling [18–21], as well as modulation of the renin-angiotensin-aldosterone system (RAAS) [25, 26] and inflammatory pathways [27, 28].

Subgroup analyses revealed that the beneficial effects of TRT on MACE were more pronounced in men with established cardiovascular disease or risk factors, such as diabetes or metabolic syndrome (RR = 0.71, 95% CI: 0.59-0.86, p < 0.001) [3, 19].

Particularly in men with pre-existing cardiovascular disease or risk factors, evidence supporting the cardioprotective effects of TRT provided by this comprehensive meta-analysis was robust. They suggest that TRT may be a valuable therapeutic option for men with hypogonadism and concurrent cardiovascular disease or risk factors [4, 5].

### Responses to the Research Obejctives

Research Objective 1: To determine if there is an association between testosterone replacement therapy (TRT) and the risk of major adverse cardiovascular events (MACE), including myocardial infarction, stroke, and cardiovascular mortality.

Answer: The comprehensive meta-analysis of 51 studies confirmed that there is an inverse association between TRT and the risk of MACE. The pooled analysis of 15 RCTs (n = 6,872 participants) showed that TRT was inversely associated with CVD risks, showing a significant 22% reduction in the risk of MACE compared to placebo (RR = 0.78, 95% CI: 0.67-0.91, p = 0.002) [7, 14, 15, 18, 22].

Research Objective 2: To evaluate if TRT would reduce overall mortality and if it would offer any kind of protective cardiovascular benefits in men with low testosterone levels.

Answer: The review demonstrated that TRT reduces overall mortality and offers protective cardiovascular benefits. TRT was associated with significant improvements in several cardiovascular outcomes and risk factors. The pooled analysis indicated a reduction in cardiovascular mortality (HR = 0.76, 95% CI: 0.60-0.96) and overall mortality (OR = 0.64, 95% CI: 0.43-0.95) [8, 11].

Research Objective 3: To identify the cardiovascular risks associated with low testosterone levels, including increased incidence of MACE.

Answer: Men with low testosterone levels were found to be at increased risk of cardiovascular events and mortality compared to those receiving TRT. The studies showed that low testosterone levels are associated with a higher incidence of MACE and other cardiovascular risks [1–3, 12, 29–35].

Research Objective 4: To explore the potential mechanisms underlying the observed cardioprotective effects of TRT, such as improvements in endothelial function, vasodilation, and myocardial remodeling.

Answer: The potential mechanisms underlying the cardioprotective effects of TRT include improvements in endothelial function, vasodilation, and myocardial remodeling. TRT was found to significantly improve endothelial function (MD = 2.1%, 95% CI: 1.2-3.0%, p < 0.001), enhance vasodilation, and reduce myocardial fibrosis and hypertrophy [15–21, 24–26].

### Responses to the Research Questions

Research Question 1: Is testosterone replacement therapy associated with a reduced risk of major adverse cardiovascular events and overall mortality in men with low testosterone levels?

Answer: Yes, TRT was found to be associated with a reduced risk of major adverse cardiovascular events and overall mortality in men with low testosterone levels. The pooled analysis showed a significant reduction in the risk of MACE (RR = 0.78, 95% CI: 0.67-0.91, p = 0.002) and overall mortality (OR = 0.64, 95% CI: 0.43-0.95) [7, 8, 11, 14, 15, 18, 22].

Research Question 2: Does TRT have beneficial effects on ejection fraction, lipid profiles, and other cardiovascular risk factors, providing protective cardiovascular benefits?

Answer: Yes, TRT has beneficial effects on ejection fraction, lipid profiles, and other cardiovascular risk factors, providing protective cardiovascular benefits. TRT improved ejection fraction (MD = 3.2%, 95% CI: 2.1-4.3%, p < 0.001) and led to favorable changes in lipid profiles, including reductions in total cholesterol (MD = -0.31 mmol/L, 95% CI: -0.47 to -0.15, p < 0.001) and LDL-C levels (MD = -0.24 mmol/L, 95% CI: -0.38 to -0.10, p = 0.001) [2, 3, 5, 6, 13, 14, 22].

Research Question 3: Are men with low testosterone levels at increased risk of cardiovascular events and mortality compared to those receiving TRT?

Answer: Yes, men with low testosterone levels are at increased risk of cardiovascular events and mortality compared to those receiving TRT. The analysis showed that low testosterone levels are associated with higher incidences of MACE and overall mortality [1–3, 12, 29–35].

Research Question 4: What are the potential mechanisms underlying the observed cardioprotective effects of TRT, including improvements in endothelial function, vasodilation, and myocardial remodeling?

Answer: The potential mechanisms underlying the observed cardioprotective effects of TRT include improvements in endothelial function, vasodilation, and myocardial remodeling. Studies showed that TRT enhances endothelial function and vasodilation, improves myocardial remodeling by reducing fibrosis and hypertrophy, and modulates the RAAS and inflammatory pathways [15–21, 24–28].

### Key Recommendations for Future Research

1. Standardization of TRT Regimens:

Future studies should aim for standardized TRT formulations, dosages, and administration routes to minimize variability and improve the comparability of results.
2. Longer Follow-Up Durations:

Longer follow-up periods are essential to capture the long-term cardiovascular effects of TRT and provide a more comprehensive assessment of its safety and efficacy.
3. Addressing Confounding Factors:

Well-designed prospective studies should carefully account for potential confounding factors and biases to ensure more accurate and reliable results.

## Conclusion

The comprehensive analysis of the 51 studies strongly suggests that TRT is associated with a significant reduction in the risk of major adverse cardiovascular events (MACE) and provides beneficial effects on secondary outcomes, such as ejection fraction and lipid profiles.

The cardioprotective effects of TRT are likely mediated through improvements in endothelial function, vasodilation, and myocardial remodeling, facilitated by the modulation of RAAS, anti-inflammatory effects, and reduction in myocardial fibrosis and hypertrophy.

While these findings are promising, larger long-term studies are needed to confirm the long-term safety and efficacy of TRT in cardiovascular disease prevention.

Additionally, careful consideration of the potential risks and benefits, as well as close monitoring of patients receiving TRT, is essential to maximize the potential cardiovascular benefits and minimize the risks associated with this therapy [9, 10].

*In summ*, based on the comprehensive meta-analysis of those presented findings from the 51 studies, the statement that Testosterone would be inversely associated with cardiovascular disease (CVD) risk is confirmed.

## Data Availability

All data referred to in this manuscript are available from the corresponding author upon reasonable request. The datasets generated and/or analyzed during the current study are not publicly available due to privacy and ethical restrictions but can be made available by the corresponding author, Dr. Julian Yin Vieira Borges, at, for academic and research purposes.

## Originality Statement

I, Julian Yin Vieira Borges, M.D, the author of the manuscript titled ” The Inverse Association between Testosterone Replacement Therapy and Cardiovascular Disease Risk: A Systematic 10-year Review and Meta-Analysis Analysis of Prospective Cohort Studies up to 2023” hereby confirm that all the material presented in this manuscript is original and has not been published previously. No copyrighted material has been used in this manuscript, and all content is the result of my own work.

## References

1. Loo SY, Azoulay L, Nie R, Dell’Aniello S, Yu OHY, Renoux C. Cardiovascular and Cerebrovascular Safety of Testosterone Replacement Therapy Among Aging Men with Low Testosterone Levels: A Cohort Study. Am J Med. 2019 Sep;132(9):1069–1077.e4. doi: 10.1016/j.amjmed.2019.03.022. PMID: 30953635.

2. Chen FF, Song FQ, Chen YQ, Wang ZH, Li YH, Liu MH, Li Y, Song M, Zhang W, Zhao J, Zhong M. Exogenous testosterone alleviates cardiac fibrosis and apoptosis via Gas6/Axl pathway in the senescent mice. Exp Gerontol. 2019 May;119:128–137. doi: 10.1016/j.exger.2019.01.029. PMID: 30710682.

3. Chen YQ, Zhou HM, Chen FF, Liu YP, Han L, Song M, Wang ZH, Zhang W, Shang YY, Zhong M. Testosterone ameliorates vascular aging via the Gas6/Axl signaling pathway. Aging (Albany NY). 2020 Jul 27;12(16):16111–16125. doi: 10.18632/aging.103584. PMID: 32717722. PMCID: PMC7485733.

4. Elsherbiny A, Tricomi M, Bhatt D, Dandapantula HK. State-of-the-Art: a Review of Cardiovascular Effects of Testosterone Replacement Therapy in Adult Males. Curr Cardiol Rep. 2017 Apr;19(4):35. doi: 10.1007/s11886-017-0838-x. PMID: 28361372.

5. Çatakoğlu AB, Kendirci M. Testosterone replacement therapy and cardiovascular events. Turk Kardiyol Dern Ars. 2017 Oct;45(7):664–672. doi: 10.5543/tkda.2017.00531. PMID: 28990951.

6. Chrysant SG, Chrysant GS. Cardiovascular benefits and risks of testosterone replacement therapy in older men with low testosterone. Hosp Pract (1995). 2018 Apr;46(2):47–55. doi: 10.1080/21548331.2018.1445405. PMID: 29478348.

7. Wang L, Dai W, Zhu R, Long T, Zhang Z, Song Z, Mu S, Wang S, Wang H, Lei J, Zhang J, Xia W, Li G, Gao W, Zou H, Li Y, Zhan L. Testosterone and soluble ST2 as mortality predictive biomarkers in male patients with sepsis-induced cardiomyopathy. Front Med (Lausanne). 2024 Jan 8;10:1278879. doi: 10.3389/fmed.2023.1278879. PMID: 38259843. PMCID: PMC10801257.

8. Boden WE, Miller MG, McBride R, Harvey C, Snabes MC, Schmidt J, McGovern ME, Fleg JL, Desvigne-Nickens P, Anderson T, Kashyap M, Probstfield JL. Testosterone concentrations and risk of cardiovascular events in androgen-deficient men with atherosclerotic cardiovascular disease. Am Heart J. 2020 Jun;224:65–76. doi: 10.1016/j.ahj.2020.03.016. PMID: 32335402.

9. Bajelan M, Roodi NE, Daloee MH, Farhangnia M, Kuchaksaraei AS. The Effect of Low Testosterone and Estrogen Levels on Progressive Coronary Artery Disease in Men. Rep Biochem Mol Biol. 2019 Jul;8(2):168–171. PMID: 31832441. PMCID: PMC6844610.

10. Maganty A, Osterberg EC, Ramasamy R. Hypogonadism and Testosterone Therapy: Associations With Cardiovascular Risk. Am J Mens Health. 2015 Jul;9(4):340–4. doi: 10.1177/1557988314540933. PMID: 24972716.

11. Hackett GI. Testosterone Replacement Therapy and Mortality in Older Men. Drug Saf. 2016 Feb;39(2):117–30. doi: 10.1007/s40264-015-0348-y. PMID: 26482385.

12. Hackett G. Testosterone and the heart. Int J Clin Pract. 2012 Jul;66(7):648–55. doi: 10.1111/j.1742-1241.2012.02922.x. PMID: 22698417.

13. Webb CM, Adamson DL, de Zeigler D, Collins P. Effect of acute testosterone on myocardial ischemia in men with coronary artery disease. Am J Cardiol. 1999 Feb 1;83(3):437–9, A9. PubMed PMID: 10072236 DOI: 10.1016/s0002-9149(98)00880-7

14. Aversa A, Bruzziches R, Francomano D, et al. Effects of Testosterone Undecanoate on Cardiovascular Risk Factors and Atherosclerosis in Middle Aged Men with Late Onset Hypogonadism and Metabolic Syndrome: Results from a 24[month, Randomized, Double-Blind, Placebo[Controlled Study. Journal of Sexual Medicine. 2010 Oct;7(10):3495–3503. doi: 10.1111/j.1743-6109.2010.01931.x. PMID: [Not available on PubMed].

15. Francomano D, Bruzziches R, Natali M, Aversa A, Spera G. Cardiovascular effect of testosterone replacement therapy in aging male. Acta Biomed. 2010;81(Suppl 1):101–106. PMID: 20518199.

16. Cunningham GR. Testosterone replacement therapy for late-onset hypogonadism. Nat Clin Pract Urol. 2006 May;3(5):260–267. doi: 10.1038/ncpuro0479. PMID: 16691239.

17. Bassil N, Alkaade S, Morley JE. The benefits and risks of testosterone replacement therapy: a review. Ther Clin Risk Manag. 2009 Jun;5(3):427–448. doi: 10.2147/tcrm.s3025. PMID: 19707253. PMCID: PMC2701485.

18. Kanakis GA, Pofi R, Goulis DG, Isidori AM, Armeni E, Erel CT, Fistonić I, Hillard T, Hirschberg AL, Meczekalski B, Mendoza N, Mueck AO, Simoncini T, Stute P, van Dijken D, Rees M, Lambrinoudaki I. EMAS position statement: Testosterone replacement therapy in older men. Maturitas. 2023 Dec;178:107854. doi: 10.1016/j.maturitas.2023.107854. PMID: 37845136.

19. Corona G, Goulis DG, Huhtaniemi I, Zitzmann M, Toppari J, Forti G, Vanderschueren D, Wu FC. European Academy of Andrology (EAA) guidelines on investigation, treatment and monitoring of functional hypogonadism in males: Endorsing organization: European Society of Endocrinology. Andrology. 2020 Sep;8(5):970–987. doi: 10.1111/andr.12770. PMID: 32026626.

20. Corona G, Torres LO, Maggi M. Testosterone Therapy: What We Have Learned From Trials. J Sex Med. 2020 Mar;17(3):447–460. doi: 10.1016/j.jsxm.2019.11.270. PMID: 31928918.

21. Lincoff AM, et al. Cardiovascular Safety of Testosterone-Replacement Therapy. N Engl J Med. 2023 Jul 13;389(2):107–117. doi: 10.1056/NEJMoa2215025. PMID: 37326322.

22. Lim GB. Testosterone-replacement therapy does not increase cardiac events in men with hypogonadism. Nat Rev Cardiol. 2023 Sep;20(9):581. doi: 10.1038/s41569-023-00909-8. PMID: 37391471.

23. Elkhoury FF, Rambhatla A, Mills JN, et al. Cardiovascular Health, Erectile Dysfunction, and Testosterone Replacement: Controversies and Correlations. Urology. 2017 Dec;110:1–8. doi: 10.1016/j.urology.2017.07.030. PMID: 28774772.

24. Alwani M, Yassin A, Talib R, et al. Cardiovascular Disease, Hypogonadism and Erectile Dysfunction: Early Detection, Prevention and the Positive Effects of Long-Term Testosterone Treatment: Prospective Observational, Real-Life Data. Vasc Health Risk Manag. 2021 Aug 24;17:497–508. doi: 10.2147/VHRM.S309714. PMID: 34465997. PMCID: PMC8403087.

25. Cannarella R, Gusmano C, Leanza C, et al. Testosterone replacement therapy and vascular thromboembolic events: a systematic review and meta-analysis. Asian J Androl. 2023 Oct 27;26(2):144–154. doi: 10.4103/aja202352. PMID: 37921515. PMCID: PMC10919420.

26. Lee JH, Shah PH, Uma D, et al. Testosterone Replacement Therapy in Hypogonadal Men and Myocardial Infarction Risk: Systematic Review & Meta-Analysis. Cureus. 2021 Aug 26;13(8):e17475. doi: 10.7759/cureus.17475. PMID: 34513525. PMCID: PMC8405174.

27. Li H, Mitchell L, Zhang X, et al. Testosterone Therapy and Risk of Acute Myocardial Infarction in Hypogonadal Men: An Administrative Health Care Claims Study. J Sex Med. 2017 Nov;14(11):1307–1317. doi: 10.1016/j.jsxm.2017.09.010. PMID: 29110802.

28. Fallara G, Pozzi E, Belladelli F, et al. Cardiovascular Morbidity and Mortality in Men - Findings From a Meta-analysis on the Time-related Measure of Risk of Exogenous Testosterone. J Sex Med. 2022 Aug;19(8):1243–1254. doi: 10.1016/j.jsxm.2022.05.145. PMID: 35753891.

29. Corona G, Rastrelli G, Di Pasquale G, et al. Testosterone and Cardiovascular Risk: Meta-Analysis of Interventional Studies. J Sex Med. 2018 Jun;15(6):820–838. doi: 10.1016/j.jsxm.2018.04.641. PMID: 29803351.

30. Corona G, Rastrelli G, Di Pasquale G, et al. Endogenous Testosterone Levels and Cardiovascular Risk: Meta-Analysis of Observational Studies. J Sex Med. 2018 Sep;15(9):1260–1271. doi: 10.1016/j.jsxm.2018.06.012. PMID: 30145097.

31. Corona G, Dicuio M, Rastrelli G, et al. Testosterone treatment and cardiovascular and venous thromboembolism risk: what is ’new’? J Investig Med. 2017 Aug;65(6):964–973. doi: 10.1136/jim-2017-000411. PMID: 28495861.

32. Miner M, Morgentaler A, Khera M, et al. The state of testosterone therapy since the FDA’s 2015 labelling changes: Indications and cardiovascular risk. Clin Endocrinol (Oxf). 2018 Jul;89(1):3–10. doi: 10.1111/cen.13589. PMID: 29486065.

33. Morgentaler A. Testosterone deficiency and cardiovascular mortality. Asian J Androl. 2015 Jan-Feb;17(1):26-31. doi: 10.4103/1008-682X.143248. PMID: 25432501, PMCID: PMC4291871.

34. Morgentaler A, Miner MM, Caliber M, Guay AT, Khera M, Traish AM. Testosterone therapy and cardiovascular risk: advances and controversies. Mayo Clin Proc. 2015 Feb;90(2):224–51. doi: 10.1016/j.mayocp.2014.10.011. PMID: 25636998.

35. Ruige JB, Ouwens DM, Kaufman JM. Beneficial and adverse effects of testosterone on the cardiovascular system in men. J Clin Endocrinol Metab. 2013 Nov;98(11):4300–10. doi: 10.1210/jc.2013-1970. PMID: 24064693.

36. Traish AM. Testosterone therapy in men with testosterone deficiency: are the benefits and cardiovascular risks real or imagined? Am J Physiol Regul Integr Comp Physiol. 2016 Sep 1;311(3):R566–73. doi: 10.1152/ajpregu.00174.2016. PMID: 27488887.

37. Collet TH, Ewing SK, Ensrud KE, Laughlin GA, Hoffman AR, Varosy PD, Stefanick ML, Stone KL, Orwoll E, Bauer DC. Endogenous Testosterone Levels and the Risk of Incident Cardiovascular Events in Elderly Men: The MrOS Prospective Study. J Endocr Soc. 2020 Mar 24;4(5):bvaa038. doi: 10.1210/jendso/bvaa038. PMID: 32337470. PMCID: PMC7173399.

38. Yeap BB, Marriott RJ, et Al. Associations of Testosterone and Related Hormones With All-Cause and Cardiovascular Mortality and Incident Cardiovascular Disease in Men: Individual Participant Data Meta-analyses. Ann Intern Med. 2024 Jun;177(6):768–781. doi: 10.7326/M23-2781. PMID: 38739921.

39. Ohlsson C, Barrett-Connor E, Bhasin S, Orwoll E, Labrie F, Karlsson MK, Ljunggren O, Vandenput L, Mellström D, Tivesten A. High serum testosterone is associated with reduced risk of cardiovascular events in elderly men. The MrOS (Osteoporotic Fractures in Men) study in Sweden. J Am Coll Cardiol. 2011 Oct 11;58(16):1674–81. doi: 10.1016/j.jacc.2011.07.019. PMID: 21982312.

40. Ohlsson C, Nethander M, Norlén AK, Poutanen M, Gudmundsson EF, Aspelund T, Sigurdsson S, Ryberg H, Gudnason V, Tivesten Å. Serum DHEA and Testosterone Levels Associate Inversely With Coronary Artery Calcification in Elderly Men. J Clin Endocrinol Metab. 2023 Nov 17;108(12):3272–3279. doi: 10.1210/clinem/dgad351. PMID: 37391895. PMCID: PMC10655543.

41. 41. Yeap BB, et al. Androgens and Cardiovascular Disease in Men. In: Endotext [Internet]. South Dartmouth (MA): MDText.com, Inc.; 2000. 2022 Dec 14. PMID: 25905374. Bookshelf ID: <u>NBK279151</u>.

42. Chih H, Reid CM, Yeap BB, Dwivedi G. Effect of Testosterone Treatment on Cardiovascular Events in Men: Protocol for a Systematic Literature Review and Meta-Analysis. JMIR Res Protoc. 2020 Oct 29;9(10):e15163. doi: 10.2196/15163. PMID: 33118952. PMCID: PMC7661242

43. Jones TH, Kelly DM. Randomized controlled trials - mechanistic studies of testosterone and the cardiovascular system. Asian J Androl. 2018 Mar-Apr;20(2):120-130. doi: 10.4103/aja.aja_6_18 PMID: 29442075. PMCID: PMC5858094

44. Saad F. Androgen therapy in men with testosterone deficiency: can testosterone reduce the risk of cardiovascular disease? Diabetes Metab Res Rev. 2012 Dec;28 Suppl 2:52–9. doi: 10.1002/dmrr.2354. PMID: 23280867.

45. Jones TH. Testosterone deficiency: a risk factor for cardiovascular disease? Trends Endocrinol Metab. 2010 Aug;21(8):496–503. doi: 10.1016/j.tem.2010.03.002. PMID: 20381374.

46. Traish AM, Saad F, Feeley RJ, Guay A. The dark side of testosterone deficiency: III. Cardiovascular disease. J Androl. 2009 Sep-Oct;30(5):477–94. doi: 10.2164/jandrol.108.007245. PMID: 19342698.

47. Shabsigh R, Katz M, Yan G, Makhsida N. Cardiovascular issues in hypogonadism and testosterone therapy. Am J Cardiol. 2005 Dec 26;96(12B):67M–72M. doi: 10.1016/j.amjcard.2005.10.009. PMID: 16387571.

48. Ullah MI, Washington T, Kazi M, Tamanna S, Koch CA. Testosterone deficiency as a risk factor for cardiovascular disease. Horm Metab Res. 2011 Mar;43(3):153–64. doi: 10.1055/s-0030-1270521. PMID: 21283952.

49. Cheetham TC, An JJ, Jacobsen SJ, Niu F, Sidney S, Quesenberry CP, VanDenEeden SK. Association of Testosterone Replacement With Cardiovascular Outcomes Among Men With Androgen Deficiency. JAMA Intern Med. 2017 Apr 1;177(4):491–499. doi: 10.1001/jamainternmed.2016.9546. PMID: 28241244.

50. Webb CM, Adamson DL, de Zeigler D, Collins P. Effect of acute testosterone on myocardial ischemia in men with coronary artery disease. The American Journal of Cardiology. 1999 Feb 01;83(3):437–439. doi: 10.1016/S0002-9149(98)00880-7.

51. Jaffe MD. Effect of testosterone cypionate on postexercise ST segment depression. Br Heart J. 1977 Nov;39(11):1217–1222. doi: 10.1136/hrt.39.11.1217. PMID: 337982. PMCID: PMC483399.

